# The Effects of Metronome Frequency Differentially Affects Gait on a Treadmill and Overground in People with Parkinson’s Disease

**DOI:** 10.1101/19003251

**Authors:** Madelon Wygand, Guneet Chawla, Nina Browner, Michael D Lewek

## Abstract

**Objective:** To determine the effect of different metronome cue frequencies on spatiotemporal gait parameters when walking overground compared to walking on a treadmill in people with Parkinson’s disease

**Design:** Repeated-measures, within-subject design

**Setting:** Research laboratory

**Participants:** Twenty-one people with Parkinson’s disease (Hoehn & Yahr stage 1-3)

**Interventions:** Participants walked overground and on a treadmill with and without metronome cues of 85%, 100%, and 115% of their baseline cadence for one minute each.

**Main Outcome Measures:** Gait speed, step length, and cadence

**Results:** An interaction effect between cue frequency and walking environment revealed that participants took longer steps during the 85% condition on the treadmill only. When walking overground, metronome cues of 85% and 115% of baseline cadence yielded decreases and increases, respectively, in both cadence and gait speed with no concomitant change in step length.

**Conclusions:** These data suggest that people with PD are able to alter spatiotemporal gait parameters immediately when provided the appropriate metronome cue and walking environment. We propose to target shortened step lengths by stepping to the beat of slow frequency auditory cues while walking on a treadmill, whereas the use of fast frequency cues during overground walking can facilitate faster walking speeds.

---

Common gait alterations for individuals with Parkinson’s disease (PD) include decreased stride length, increased variability of movement, freezing of gait, decreased gait speed, increased double support time, and increased cadence.^1-5^ These gait abnormalities are associated with decreased motor automaticity which begins to deteriorate in the early stages of PD.^6^ In an effort to improve gait mechanics and quality of life for individuals with PD, physical therapy often targets the rhythmicity of gait.^7^

Walking on a motorized treadmill can encourage more automatic gait for individuals with PD, due to the continuous belt movement compared to the self-generating gait pattern performed overground.^8, 9^ As a result, walking on a treadmill leads to decreased gait variability when compared to overground gait.^10^ Even a single session of treadmill walking can elicit longer steps for individuals with PD.^11^ This may be because individuals with advanced PD, unlike unimpaired individuals, may take increased step lengths and decrease the cadence when walking on a treadmill compared to walking overground.^12^ These observed changes in gait following treadmill walking might be due to a combination of the proprioceptive input from the belt displacement of the limbs, visual cues of the distance from the front of the treadmill, or the constant velocity of the belt.^10, 12^

The use of rhythmic auditory cues has likewise successfully improved gait parameters in people with PD.^13, 14^ Rhythmic auditory cueing presumably serves as an external auditory stimulus to bypass the impaired internal timing present in PD.^15, 16^ Generally, the use of rhythmic auditory cues has contributed to increases in gait speed and step lengths,^13, 17, 18^ although this finding is inconsistent.^5, 19^ Importantly, however, the prescriptive literature has been narrowly focused on the use of *faster* tempos of rhythmic auditory cueing.^14^ This is presumably because most investigators primarily seek to increase gait velocity, and therefore suggest that cueing should only be used at frequencies that match or exceed the individual’s baseline cadence.^15, 20^ Furthermore, the use of slower cueing frequencies has been suggested to increase falls risk.^20^

Of concern is that auditory cueing is intended to impact the temporal features of gait,^21^ but impacting step length may have a greater influence on gait for individuals with PD.^5, 22^ Gait speed is a function of both temporal (e.g., cadence) and spatial (e.g., step length) features.^23^ Therefore, auditory cue-induced changes in cadence when walking overground may increase gait speed, but without obliging an individual to increase step length. Furthermore, given the potentially valuable role of walking on a treadmill for individuals with PD, ^8, 10, 24^ we are concerned that increasing cadence with rhythmic auditory cueing while walking at a constant treadmill speed will induce even shorter steps. Clearly, the environmental context of rhythmic auditory cueing matters. Therefore, the purpose of this study was to determine the effect of metronome cues on spatiotemporal gait parameters when walking overground compared to walking on a treadmill in people with PD. This information will be critical in order to determine the optimal training environment to achieve specific gait training goals. We hypothesize that individuals with PD will increase step length with *slow* tempo metronome cueing during treadmill walking due to the inability to change gait speed, but will increase step length overground while stepping to *fast* tempo metronome cueing.

## Methodology

### Participants

We recruited participants from the community via referrals from the Department of Neurology and local PD support groups. All participants were diagnosed with PD by a physician, and had a Hoehn and Yahr (H&Y) Stage 1-3. Additional inclusion criteria included self-reported ability to walk >10 m over ground and on a treadmill for at least 14 minutes with rest breaks as needed. Exclusion criteria included H&Y Stage 4 and 5, uncontrolled cardiorespiratory/metabolic disease, other neurologic/orthopedic disorders that affect gait, and severe communication impairments that impeded understanding of study purpose and procedures. All participants signed an informed consent form approved by the UNC-Chapel Hill IRB. The project was listed on ClinicalTrials.gov (NCT03253965).

### Procedures

Participants completed the Montreal Cognitive Assessment (MoCA) and the Unified Parkinson’s Disease Rating Scale (UPDRS) prior to testing. To determine baseline gait speed, each participant walked over a 20 ft pressure mat ^a^ with instruction to walk at their “comfortable speed.” From this baseline testing, software ^b^ calculated baseline gait speed and cadence for each participant, which was used to determine the treadmill speed and metronome frequencies for the study.

Each participant performed four different walking conditions on both an instrumented treadmill and overground, in a laboratory setting. These conditions consisted of a Control (no metronome) and three metronome conditions of 85%, 100%, and 115% of the cadence from the baseline overground testing. The Control (no cue) condition was always performed first and the order of the metronome conditions was randomized using a random number generator run by the investigators. The treadmill and overground tests were counterbalanced. We played the metronome ^c^ through a speaker for all metronome conditions.

#### Overground Testing

Each participant walked over the 20 ft pressure mat for four passes of continuous walking for each condition. No assistive devices were used. Instructions were given to “match each step to the beat of the metronome.” The initial steps of each trial and the turns between each pass were completed off the walkway and were not included in the data collection or analysis. Standing rest breaks were given as needed between conditions.

#### Treadmill Testing

Participants walked on a dual-belt instrumented treadmill ^d^ for all treadmill conditions. The treadmill speed was set to the gait speed from the initial baseline overground test, and maintained for all conditions. For three subjects, the treadmill speed was reduced due to an inability to reach the intended treadmill speed safely. Participants were discouraged from using handrails to promote more natural gait mechanics, however, 12 participants required handrails for balance only. For those cases, handrail use was maintained consistently across conditions. All participants wore a harness attached overhead, but no body weight support was provided and the harness did not restrict limb motion. Oxygen saturation and heart rate were monitored for safety. Rest breaks were provided as needed.

Instructions were given to “match each step to the beat of the metronome.” Participants were given ∼15 seconds to synchronize with the metronome each time the frequency was changed, and data was then collected for one minute. As participants walked on the treadmill, the positions of the feet were tracked using retro-reflective markers placed on both heels by an 8-camera motion capture system ^e^ at 120 Hz. Ground reaction force data were sampled simultaneously at 1200 Hz from the treadmill. Results of the treadmill testing make up a portion of another report.^25^

### Data Management

For all overground trials, we used the PKMAS software to remove partial steps and calculate average step length, cadence, and gait speed from all remaining steps. Data were processed separately for each condition. For all treadmill trials, data were analyzed with custom Labview ^f^ programs. In particular, step length was measured as the anteroposterior distance between heel markers at heel strike. Step time was calculated as the time, in seconds, between successive heel strikes. Cadence was then calculated as the inverse of step time. We determined cadence accuracy as the difference between the actual cadence and the intended cadence.

### Data Analysis

We used SPSS ^g^ to perform separate two-way repeated measures ANOVAs (repeated for metronome condition and walking environment) for step length, cadence, and cadence accuracy. If significant effects were found, one-way ANOVAs or paired sample t-tests were used, as appropriate. A Bonferroni correction was used to account for multiple comparisons. Because gait speed did not change on the treadmill, we used a one-way ANOVA to assess for changes in overground gait speed. A p-value of 0.05 was used for significance. Prior to data collection, we performed a power analysis using G*Power to determine an appropriate sample size. We were unaware of any work that has compared faster and slower cue frequencies across walking environments. However, we used an effect size (f > 0.39) based on differences in step length between treadmill and overground^26^ and stride length differences between metronome cue conditions.^21^ This assumption indicated that we would need 18 participants for an alpha of 0.05 and power of 0.80.

## Results

### Participants

We contacted 27 eligible participants, of whom 21 people with PD agreed to participate (13 male, 8 female; 69.8±9.8 years old). The average MoCA score was 27±2.9. Participants had experienced symptoms for 10.5±7.7 years prior to participation. Five of the participants were classified as H&Y Stage 1, nine were classified as H&Y Stage 2, and seven were classified as H&Y Stage 3. The average UPDRS score was 19.4±13.7 with the average motor axial subscore of 4.8±3.5. Nine participants were classified as axial rigid and 12 as tremor predominant.

### Gait Speed

The treadmill speed was kept constant for all conditions (1.10±0.26 m/s), however, significant changes in gait speed were observed when stepping with different metronome frequencies during overground walking (p<0.001, η^2^_p_=0.764, Figure 1). Overground gait speed significantly increased for the 115% metronome condition (1.33±0.20 m/s; p<0.001; Cohen’s d: 1.39) and significantly decreased for the 85% metronome condition to 0.99±0.17 m/s (p=0.001; Cohen’s d: 1.07) compared to Control (no cue) walking (1.15±0.21 m/s). We did not observe any change in gait speed at the 100% metronome condition (1.19±0.19 m/s; p=0.512; Cohen’s d: 0.34).

**Figure 1:**
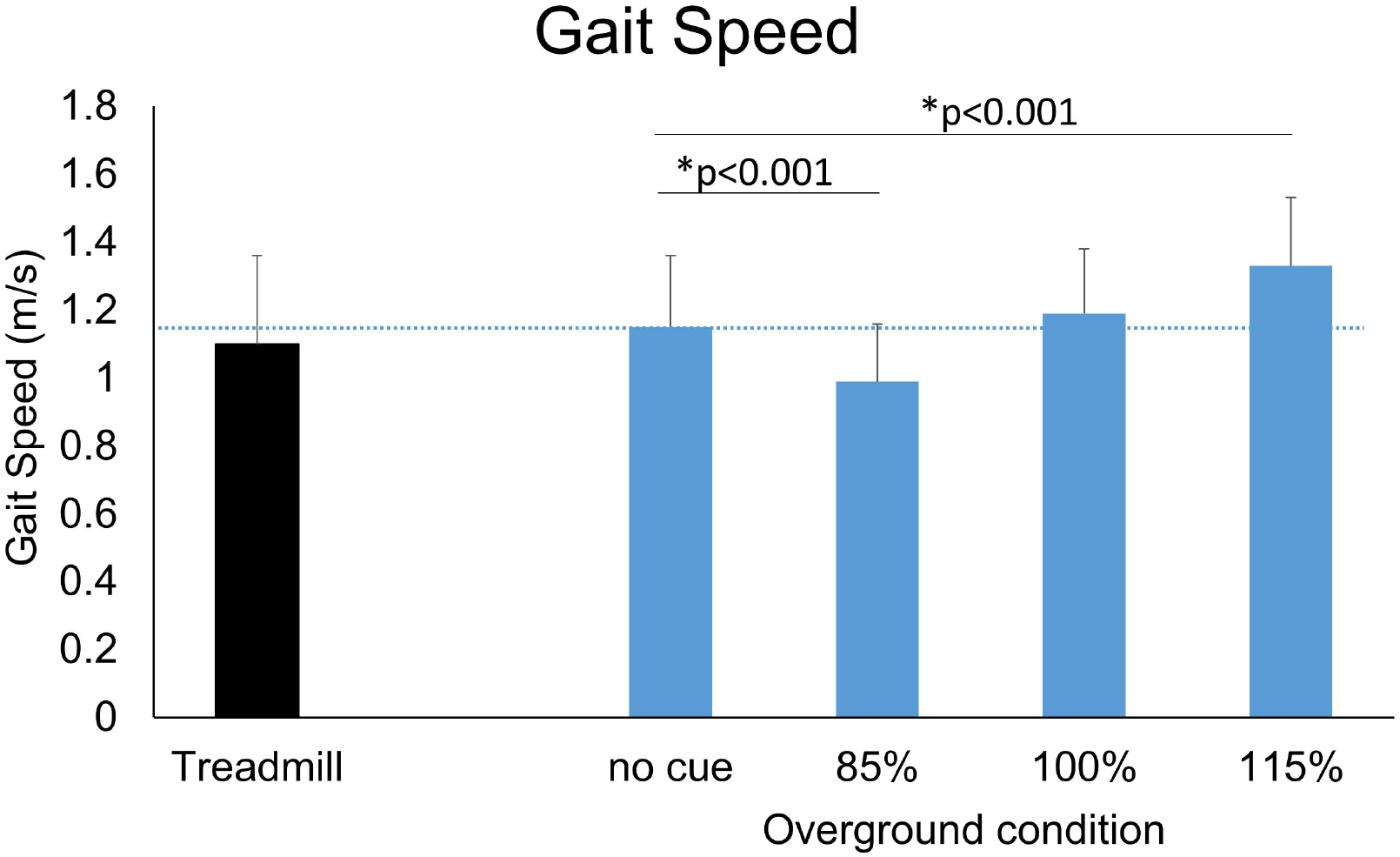
Gait speed (m/s) during overground gait demonstrated dramatic alterations with the 85% and 115% metronome conditions compared to the no cue (Control) condition. The horizontal dashed line represents the no cue condition for easier visualization of relevant comparisons. The treadmill speed did not change between conditions and is indicated on the left for reference. Error bars indicate standard deviation.

### Step Length

We observed a significant interaction effect between walking environment (treadmill vs overground) and metronome frequency (p<0.001, η^2^_p_=0.554; Figure 2). We therefore assessed the effect of metronome frequency on step length on the treadmill and overground, separately. Here, we observed that step lengths were affected by the metronome frequency on the treadmill only (p<0.001, η^2^_p_=0.761), but not during overground walking (p=0.163, η^2^_p_=0.087). Post hoc testing of treadmill walking revealed that the slower (i.e., 85%) metronome frequencies yielded significantly larger step lengths (0.61±0.13 m; p<0.001; Cohen’s d: 1.72), whereas the faster (i.e., 115%) frequencies induced smaller step lengths (0.49±0.10 m; p<0.001; Cohen’s d: 1.30) compared to treadmill walking without a metronome (0.55±0.12 m). In particular, stepping to the 85% metronome frequency induced a 10% longer step length compared to treadmill walking without auditory cues (Control).

**Figure 2:**
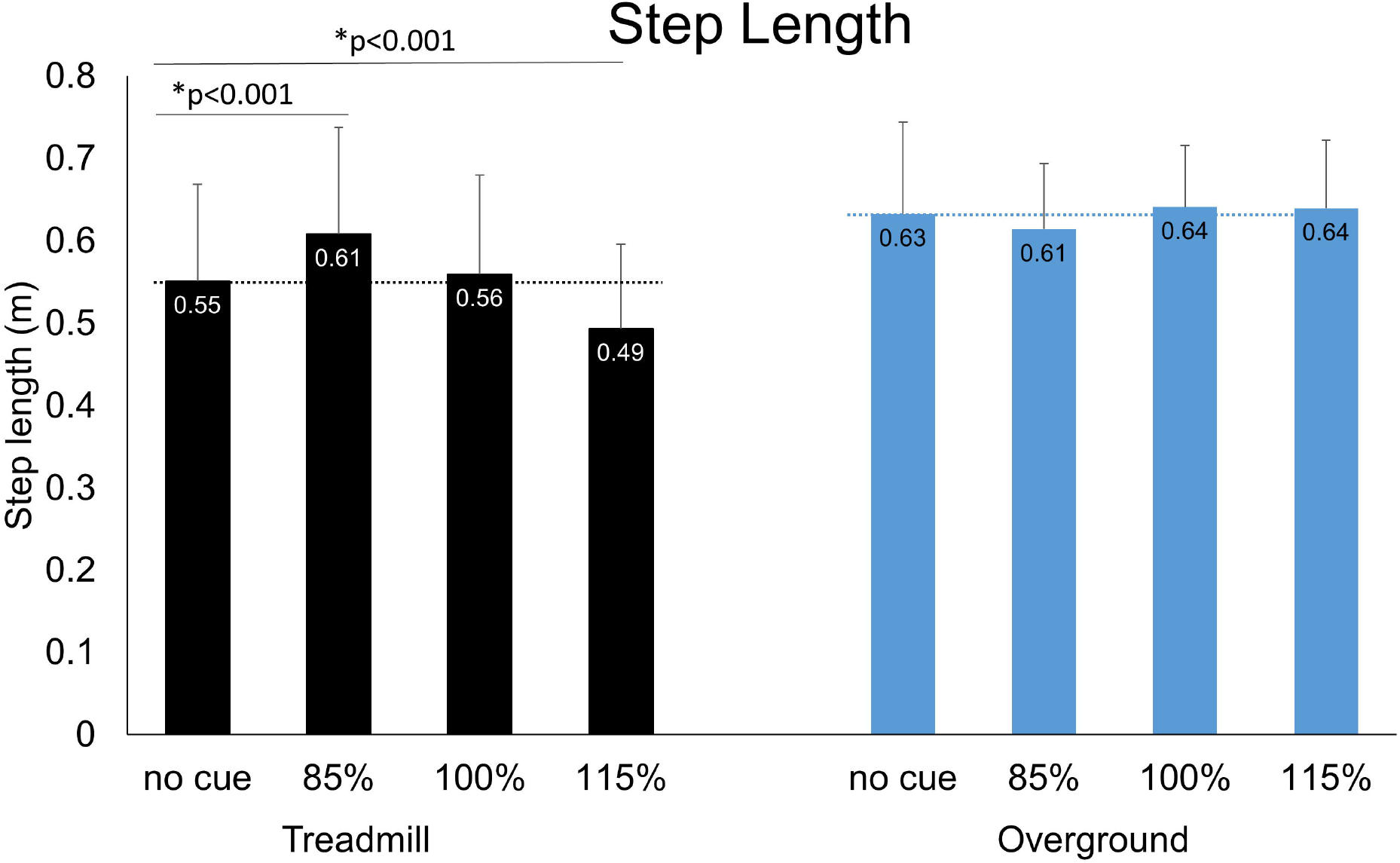
Step length (m) during treadmill gait (black) and overground gait (blue/gray). Despite no difference between conditions during overground gait, we observed significant changes during treadmill walking. Of note, the 85% condition elicited longer steps, whereas the 115% condition produced shorter steps compared to the no cue condition. The horizontal dashed line represents the respective no cue conditions for easier visualization of relevant comparisons. Error bars indicate standard deviation.

### Cadence

There was a significant interaction between walking environment and metronome frequency for cadence accuracy (p=0.014, η^2^_p_=0.224). Post-hoc evaluation revealed that participants were less accurate on the treadmill at slower (85%) tempos compared to overground (p=0.006; Cohen’s d: 0.67). Participants exhibited only small cadence errors for the 100% and 115% conditions while walking on both the treadmill and overground (p>0.256).

Cadence accuracy influenced the resulting cadence (Figure 3), as we observed an overall higher cadence on the treadmill compared to walking overground (p=0.026, η^2^_p_=0.223). Consistent with the cadence accuracy, post-hoc testing revealed that this difference in walking cadence was primarily due to the 85% condition, in which higher cadence during treadmill walking was observed compared to overground walking (p=0.006; Cohen’s d: 0.67), whereas all other frequencies yielded comparable cadences between walking environments (all p>0.256). Participants demonstrated their ability to modulate cadence (p<0.001, η^2^_p_=0.896), by slowing their cadence in the 85% condition (p<0.001; η^2^_p_=0.904) and increasing their cadence in the 115% condition (p<0.001; η^2^_p_=0.856) compared to Control walking.

**Figure 3:**
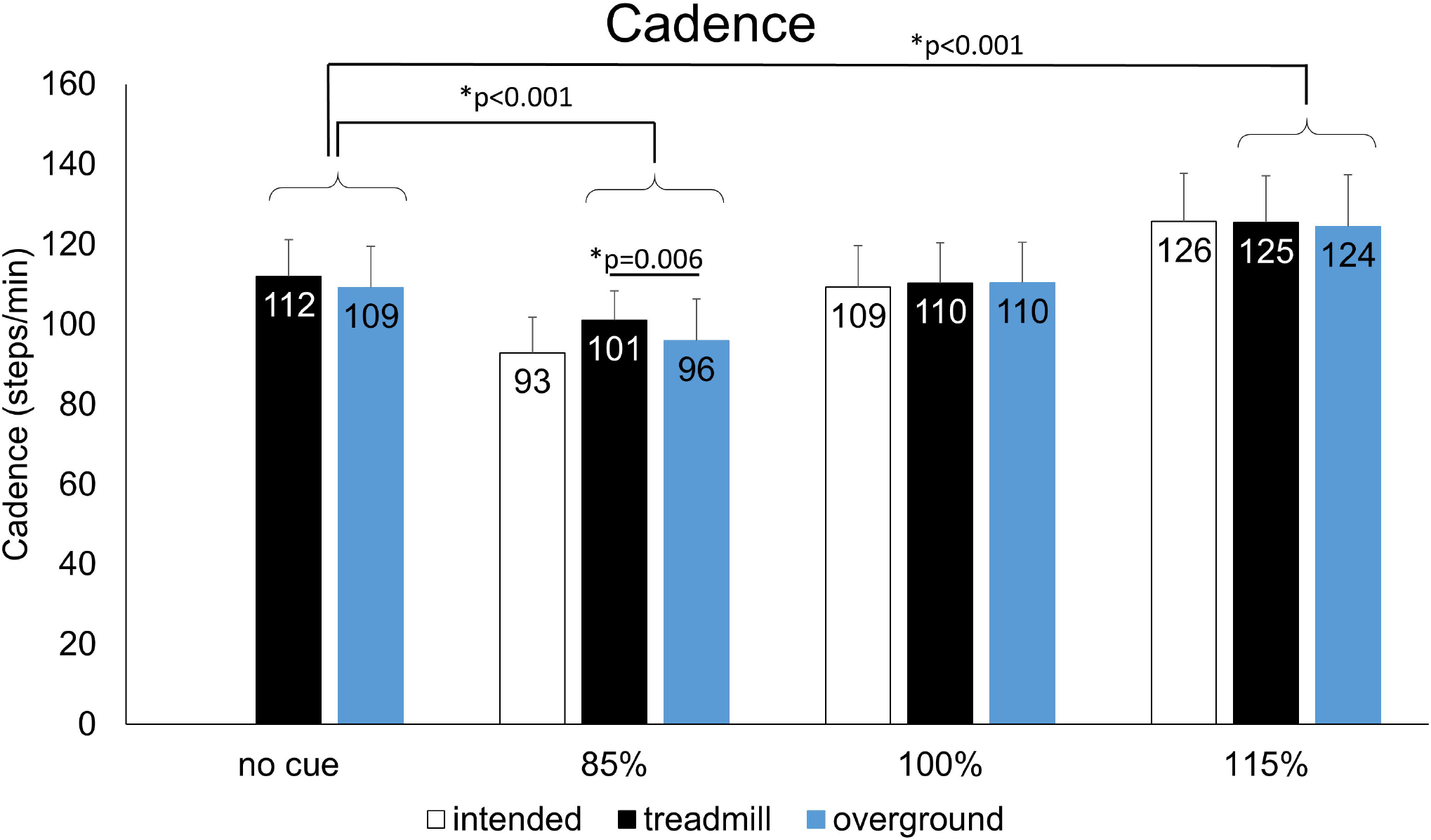
Cadence (steps/min) was influenced by the walking environment (treadmill: black; overground: blue/gray), due to the inability to match intended cadence on the treadmill during the 85% condition. Slower metronome cues reduced cadence overall, whereas faster metronome cues generated higher cadence. Examining the difference between the intended cadence and treadmill/overground values indicates the cadence accuracy. Error bars indicate standard deviation.

## Discussion

The purpose of this study was to determine the effect of metronome cue frequency on spatiotemporal gait parameters when walking overground and walking on a treadmill for individuals with PD. We had hypothesized that step length would be differentially affected by metronome frequency based on the walking environment. Indeed, we observed that step lengths were longer when walking with slow tempo cues on the treadmill only, whereas step length was not altered during overground walking. Instead, the induced change in cadence during overground walking resulted in different walking speeds without the expected alterations in step length. These results have important implications for the clinical use of rhythmic auditory cueing for individuals with PD.

These data confirm that people with PD have the ability to alter their gait immediately in response to metronome cues, but extends this finding to both overground and treadmill environments, with a particular emphasis on the specific alterations associated with different cueing frequencies. Our results likely arise from a mix of biomechanical and neuroanatomic mechanisms. In particular, the ability to accept external regulation of rhythmic timing by people with PD appears critical.^15^ Importantly, functional magnetic resonance imaging (fMRI) studies have demonstrated diminished activation in many locomotor areas of the brain in people with PD.^27^ This altered activation, in part, impairs the ability to independently modify gait mechanics through internal regulation.^28^ Use of external cues, such as the metronome and treadmill used here, places less demand on internal regulation of gait timing. Since the motor circuits for synchronization remain intact for people with PD, the use of a treadmill and rhythmic auditory cues should improve gait mechanics in ways the person would not be able to change and maintain independently.^12, 20, 21, 29^

Walking speed arises directly from spatial (e.g., step length) and temporal (e.g., cadence) measures.^23, 30^ As such, when gait speed is fixed, as on a treadmill, and cadence is manipulated, the step length must change. The same is not true for overground walking, where gait speed is free to fluctuate. Indeed, we observed predictable changes to step length on the treadmill with altered cadence, but the step lengths remained unaltered during overground walking, despite substantial alterations to cadence. These findings suggest that stepping to a *reduced* cadence tempo should reliably *increase* step lengths during treadmill walking. This approach runs counter to conventional approaches to rhythmic auditory cueing, which promote the use of faster cadences during overground walking.^15, 20^ Importantly, we did not provide our participants with any particular instruction related to altering their step length, but rather told them to simply “step with the beat”. We can speculate that if we had instructed participants to increase their step length during overground gait, that we would have seen that alteration as well. Clearly, however, the ability to modulate gait speed plays a critical role in the spatial response to cue-induced stepping.

It appears that the ability to match the cue successfully is not required to benefit from the auditory cue. Participants in this study were able to match their stepping cadence with the 100% and 115% metronome cues both overground and on the treadmill, but exhibited larger errors with the 85% metronome cue (particularly during treadmill walking). Despite the errors associated with attempting to match the 85% cues, we still observed large changes in spatiotemporal gait parameters while walking on the treadmill. Thus, even when participants did not match the cue accurately, the desired gait change was still observed.

We can use these results to guide treatment selection based on specific goals. In particular, treatment goals for individuals with PD are typically aimed at addressing common gait abnormalities, such as decreased gait speed, decreased step length, and increased cadence.^12, 28, 29^ Based on the results from our study, an immediate increase in gait speed can be realized by walking overground with the 115% metronome cue. If the treatment goal is to increase step length, we propose that walking on a treadmill with 85% metronome cues can produce a larger step length immediately. Finally, cadence is directly manipulated both on the treadmill and overground with a corresponding change to the cueing frequency. Given the specificity of the various cueing frequencies and their impact on both overground and treadmill walking, we propose a novel pairing of cue frequencies to elicit immediate changes to all relevant parameters. In particular, treadmill walking with slow metronome cues will elicit large step lengths. We propose that immediately following this “priming” stepping practice with overground walking using fast metronome cues will allow for faster walking with long step lengths. Future work is needed to determine if this novel pairing will result in substantial changes to spatiotemporal gait parameters for individuals with PD.

### Study Limitations

There were several limitations to this study. First, some participants walked slower on the treadmill compared to overground (N=3). As a result, the step lengths appeared somewhat shorter during treadmill walking compared to overground walking in our cohort. Furthermore, the procedure for computing step length was different for the treadmill and overground gait. In short, the step length from the treadmill gait likely underestimates the true step length due to trailing limb heel rise prior to leading limb heel strike. Although our procedure allows for valid within-environment comparisons, our between-environment comparisons should be interpreted with caution. Additionally, some participants used the handrails on the treadmill and some did not. This is likely because handrails provide support for improved balance and mobility.^10^ In healthy adults, there is no difference in step length between holding or not holding handrails on a treadmill.^31^ Most untrained people, especially those with PD, require handrails to perform treadmill walking safely and many studies do not report whether handrails were used. Although gait variability was not one of the outcomes assessed in this study, there is potential for differences in gait mechanics depending on use of handrails. There was also potential for fatigue-related changes in gait since rest breaks were only provided if requested, or unless heart rate or gait quality indicated the need for one. Lower extremity fatigue in PD can affect gait parameters, including step length and gait speed.^32^ Finally, we tested slightly more people with H&Y stage 2 and 3 than stage 1, suggesting that our results may be more heavily influenced by more advanced PD.

## Conclusions

In conclusion, we observed a significant relationship between the frequency of metronome cues and walking environment for gait mechanics in people with PD. The gait change that is desired (i.e. gait speed, step length, or cadence) appears to directly influences the type of cue and environment that will be needed to achieve that particular goal. People with PD are able to change their gait speed, step length, and cadence successfully in response to various cue frequencies during both overground and treadmill walking; however, further exploration is needed to determine if these results can be used as a gait training technique that carries over to long-term gait changes.

## Data Availability

Data will be made available upon request.

## List of Abbreviations

PD: Parkinson’s disease
H&Y: Hoehn and Yahr
MoCA: Montreal Cognitive Assessment
UPDRS: Unified Parkinson’s Disease Rating Scale
ANOVA: Analysis of Variance

## Supplier List

a. Zeno, Protokinetics, Havertown, PA, USA
b. PKMAS, Protokinetics, Haverton, PA, USA
c. Google metronome, Mountain View, CA, USA
d. Bertec Corp., Columbus, OH, USA
e. MX40+, Vicon, Los Angeles, CA
f. National Instruments, Austin, TX, USA
g. SPSS, ver 25, IBM, Chicago, IL, USA

## Notes

### Competing Interest Statement

Nina Browner has received travel and speaker fees from Parkinson Foundation and a grant for Parkinson Foundation Center of Excellence at University of North Carolina at Chapel Hill. For the remaining authors none were declared.

### Clinical Trial

NCT03253965

### Funding Statement

No external funding was received for this study.

### Author Declarations

All relevant ethical guidelines have been followed and any necessary IRB and/or ethics committee approvals have been obtained.

Any clinical trials involved have been registered with an ICMJE-approved registry such as ClinicalTrials.gov and the trial ID is included in the manuscript.

